# Altered neural recruitment despite dual task performance recovery in athletes with repeat concussion

**DOI:** 10.1101/2024.08.19.24312271

**Authors:** Andrew C. Hagen, Brian L Tracy, Jaclyn A. Stephens

## Abstract

Sports-related concussions (SRCs) pose significant challenges to college-aged athletes, eliciting both immediate symptoms and subacute cognitive and motor function impairment. While most symptoms and impairments resolve within weeks, athletes with repeat SRCs may experience heightened risk for prolonged recovery trajectories, future musculoskeletal injuries, and long-term neurocognitive deficits. This includes impaired dual task performance and altered neurophysiology that could persist across the lifespan and elicit future pathophysiology and neurodegeneration. Thus, it is imperative to improve our understanding of neurophysiology after SRC. This study aimed to investigate the impact of repeat SRCs on dual task performance and associated neural recruitment using functional near-infrared spectroscopy (fNIRS).

A total of 37 college-aged athletes (ages 18-24) participated in this cross-sectional observational study. Among these athletes, 20 had a history of two or more SRCs, while 17 had never sustained a SRC and served as controls. Participants completed the Neuroimaging-Compatible Dual Task Screen (NC-DTS) while fNIRS measured neural recruitment in the frontoparietal attention network and the primary motor and sensory cortices.

Behavioral analysis revealed that athletes with repeat SRCs exhibited comparable single task and dual task performance to control athletes. Additionally, dual task effects (DTE), which capture performance declines in dual tasks versus single tasks, did not significantly differ between groups. Notably, the cohort of athletes with repeat SRC in this study had a longer time since their last SRC (mean = 1.75 years) than majority of previous SRC studies. Neuroimaging results indicated altered neural recruitment patterns in athletes with multiple repeat SRCs during both single and dual tasks. Specifically, athletes with repeat SRCs demonstrated increased prefrontal cortex (PFC) activation during single motor tasks compared to controls (*P* < 0.001, *d* = 0.47). Conversely, during dual tasks, these same athletes exhibited reduced PFC activation (*P* < 0.001, *d* = 0.29) and primary motor cortex (M1) activation (*P* = 0.038, *d* = 0.16) compared to their single task activation.

These findings emphasize the complex relationship between SRC history, dual task performance, and changes in neurophysiology. While athletes with repeat SRCs demonstrate recovery in behavioral dual task performance, persistent alterations in neural recruitment patterns suggest ongoing neurophysiological changes, possibly indicating compensatory neural strategies and inefficient neural resource allocation, even beyond symptom resolution and medical clearance. Understanding the compensatory neural recruitment strategies that support behavioral performance following repeat SRCs can inform return-to-play decisions, future musculoskeletal injury risk, and the long-term impact of SRCs on neurocognitive function.

## Introduction

Concussions are prevalent in college sports, with approximately four sports-related concussions (SRC) occurring per 10,000 athlete-exposures (i.e., practice or game activities).^1^ SRC typically causes acute symptoms (e.g., headaches and nausea) along with cognitive and motor impairments, such as memory or balance problems.^2^ Many of these issues resolve in the days and weeks post-injury,^2^ but there are some aspects of performance that take longer to resolve. Specifically, dual task performance – a motor and cognitive task simultaneously – tends to resolve less quickly than single task performance.^3,4^ Additionally, markers of neurophysiology, such as neuroinflammation, altered functional connectivity of neural networks, and neurometabolic changes^5^ tend to resolve much more slowly than symptoms and behavioral performance.^6^ Indeed, researchers have documented neural changes, or differences compared to never-injured peers - for up to a year post-injury,^6,7^ but it is unknown if certain neural changes, such as changes in functional connectivity of neural networks,^7^ represent recovery or ongoing impairment.^8^

Symptomology and performance deficits from SRC also tend to resolve more slowly in athletes with repeat SRCs, especially if full recovery from the initial SRC was not achieved.^8^ Additionally, some athletes with repeat SRCs often experience enduring negative consequences, such as prolonged symptomology after a new SRC,^9^ mental health conditions,^10–12^ and sleep disturbances.^13^ Further, repeat SRCs have been linked to increased risks of long-term neurocognitive deficits, which may contribute to future pathophysiology and neurodegeneration, so deficits could potentially persist throughout an individual’s lifespan. Although studies have shown that dual task performance tends to resolve by 6 months post-injury,^3^ it is unknown if dual task performance resolves more slowly for individuals with repeat SRC. Additionally, because markers of neurophysiology are also slower to resolve overall,^6–8^ it is possible that atypical neurophysiology persists for long durations in athletes with repeat SRC.

Fortunately, due to advances in neuroimaging technology, both dual task performance and task-dependent neurophysiology (localized neural recruitment) can be evaluated simultaneously using portable functional near-infrared spectroscopy (fNIRS).^14–17^ Compared with fMRI, fNIRS is portable and less susceptible to movement artifact and, thus, can measure neural recruitment during tasks that more closely resemble the demands of sports (e.g., walking/running or throwing/catching a ball).^18^ FNIRS uses near-infrared light to detect changes in localized neural recruitment via proxy measures of oxygenated hemoglobin (HbO) and deoxygenated hemoglobin (HbR).^19^ Like fMRI, this measurement is dependent on neurovascular coupling, and quantifies changes in brain activity. Specifically, decreased HbR and increased HbO represents increased localized neural activity.^19^ However, one limitation of portable fNIRS systems is they do not provide full-head coverage, so regions of interest (ROI) must be decided *a priori.* For dual task paradigms, it is logical to evaluate superficial cortical structures that support motor performance and executive function resources, as the latter is needed for both the cognitive component and allocation of limited attention resources for the dual task.^20^ Therefore, we used the fNIRS head probe to evaluate right lateralized neural attention regions and bilateral motor and sensory cortices (see Methods for detail).

The purpose of this study was to investigate the impact of repeat SRCs on dual task performance and the associated neural recruitment in collegiate athletes. Using fNIRS, we measured task-dependent neural recruitment in the right-lateralized frontoparietal attention network and bilateral primary motor and sensory cortices while athletes with repeat SRCs and control athletes performed single and dual tasks. Given previous behavioral findings,^3^ we expected that some athletes with repeat SRC will have fully resolved dual task performance, but others may not. Additionally, although many of our athletes were greater than one-year post-SRC, we still predicted that - because they have repeat SRCs - they will demonstrate significantly different task-dependent neural recruitment, as compared to never-injured controls, across multiple ROIs, representing globally altered neural recruitment patterns.

## Materials and methods

### Study design & procedure

This was a cross-sectional observational study. Participants attended a single visit to the research laboratory which lasted 1-1.5 hours. During their visit, they completed a demographic questionnaire, a computerized concussion test (data not included here), and the Neuroimaging Compatible Dual Task Screen (NC-DTS)^21^ with simultaneous fNIRS evaluation. All study procedures were approved by the Colorado State University Institutional Review Board and all participants were oriented to the procedures and provided informed written consent.

### Participants

Seventeen control college-aged athletes and 20 college-aged athletes with a history of repeat SRCs (two or more) were recruited via flyers, word-of-mouth, email listservs, and social media advertisements. Importantly, this sample size resembles other fNIRS and dual task studies.^14,16^ Demographic information of all participants was acquired via a self-report form prior to the start of data collection. Participants were included in the control group if they were between the ages of 18–24, regularly engaged (at least four days/week) in organized sports, had no diagnosed or suspected SRC, and had no history of moderate or severe traumatic brain injury (TBI). Participants were included in the repeat SRC group if they met the same criteria for being an athlete and had at least two diagnosed SRCs and reported being cleared for participation by a medical provider. Although SRC history was acquired via self-report, athletes were required to confirm the month/year of each diagnosed SRC with the study team. In the control group, most participants were right-handed (*n* = 18/20); this was also true for the repeat SRC group, where most participants were right-handed (*n* = 16/17). This study did not exclude participants with common diagnoses or medications, such as mild depression and attention hyperactivity disorder, but the distribution of these diagnoses was balanced across the groups.

### Behavioral measure

#### Neuroimaging-compatible dual task screen (NC-DTS)

The NC-DTS was developed by our lab to evaluate the neural underpinnings of single and dual task performance,^21^ and an earlier iteration of the DTS is described elsewhere.^22^ The NC-DTS includes a lower extremity (LE) subtask and an upper extremity (UE) subtask. Each subtask includes three conditions: single motor, single cognitive, and dual task. Conditions are presented in a block design to allow for simultaneous fNIRS acquisition and measurement of averaged neural responses. Specifically, the three conditions are repeated five times in a randomized block design for a total of 15 trials per subtask. PsychoPy, a stimulus presentation software,^23^ is used to randomize trials and display trial order to a study team member who gives verbal instructions to participants. The LE and UE subtasks are described in detail here:

a. LE Subtask: The single motor condition is a 30 s obstacle walk, with foam blocks (9 in x 4 in x 6 in) placed every five m along a 15 m walkway. In this task, participants must clear each 6 in tall foam block and repeat the 15 m walk as many times as possible over a 30 s block. The primary outcome is gait speed (m/s). The single cognitive condition is a verbal fluency task, where participants must generate as many English words as possible that start with an “easy” letter (i.e., letters ‘H, D, M, A, B, F, P, T, C, S’ for which there are a wide range of words)^24^ in 30 s. In this task, the outcome measure is the number of words generated without repetitions. Finally, the dual task condition is the obstacle walk and verbal fluency task performed concurrently. The letters used for the verbal fluency task are counterbalanced between the single and dual tasks and between participants.
b. UE Subtask: The single motor condition is an alternating wall-toss task. In this task, participants stand 1.5 m away from a wall and throw and catch a tennis ball while alternating their throwing and catching hands for 30 s. The outcome measure is the number of successful catches. Catches are considered successful if the ball does not touch the floor. The single cognitive condition is a serial subtraction task where participants subtract backward by sevens from a given three-digit number which ends in ‘0’ or ‘5’. The outcome measure is the number of correct subtractions, and participants are given credit for any correct subtraction even if the previous subtraction was incorrect. Finally, the dual task condition is a combination of the wall task and serial subtractions. As in the LE subtask, the numbers used for serial subtraction task are counterbalanced between the single and dual tasks and between participants.

### Scoring and analysis of NC-DTS behavioral data

All behavioral performance was video recorded and then scored by two trained members of the research team. Detailed scoring procedures are outlined in a prior publication.^21^ Due to the block design nature of subtask administration, behavioral performance was evaluated by averaging the performance from five trial repetitions for each of three conditions in the LE and UE subtask. In the LE subtask, averaged gait speed is the single motor condition performance metric, and averaged number of words is the single cognitive condition performance metric. Similarly, averaged gait speed and averaged number of words are the dual task condition performance metrics. In the UE subtask, the averaged number of catches is the single motor condition performance metric, and the averaged number of subtractions is the single cognitive condition performance metric. Similarly, the averaged number of catches and averaged number of subtractions are the dual task condition performance metrics.

### Neuroimaging measure

#### Functional near-infrared spectroscopy (fNIRS) acquisition

The NIRSport2 (NIRx Medical Technologies) is a wearable device secured via backpack-like straps and was used to acquire fNIRS data. The NIRSport2 does not provide full head coverage. Therefore, regions of interest (ROI) were established *a priori*, and the fNIRS head probe was designed accordingly. To design the head probe, we used the fNIRS Optodes Location Decider (fOLD) toolbox^25^ in MATLAB (v. R2022b) and its AAL2 atlas to identify channel locations for our ROIs. Specifically, our ROIs included the right lateralized frontoparietal attention network^26^ and the primary and supplementary motor regions. The fNIRS head probe held 30 optodes, 15 LED sources (760 and 850 nm), and 15 detectors which created 42 channels over these ROIs. Additionally, eight short-separator detectors were placed inside the fNIRS cap to measure scalp perfusion,^27^ for a total of 50 channels. The anatomical landmarks for each channel were confirmed using AtlasViewer^28^ (Figure 1, Table 1). During acquisition, fNIRS data were wirelessly transmitted to a dedicated laptop that used Aurora software (Ver. 2021.9, NIRx Medical Technologies). Immediately before fNIRS data acquisition, a signal optimization step was completed to first calibrate the amount of light needed for each light source and then confirm that high-quality data could be acquired at each channel. Aurora software displays both light source intensities for each source, and it also indicates quality levels: ‘critical’, ‘acceptable’, or ‘excellent’ for each channel, including the short-separator channels. These indicators reflect how much light is passing through cortical tissues. An ‘excellent’ quality indicator reflects light intensity values ≥ 3 mV; ‘acceptable’ reflects values between 0.5 mV and 3 mV, and critical low threshold values are ≤ 0.5 mV. Signal quality was further evaluated using a coefficient of variance, which is the ratio between the standard deviation and the mean of the raw signal (mV) across 1.5 seconds of data acquisition following source brightness optimization. Excellent coefficient of variance values are ≤ 2.5%; acceptable values are between 2.5% and 7.4%, and critical values are ≥ 7.5%. For all participants, signal optimization and cap preparation steps (e.g. moving hair, increasing tension on optodes) were repeated until all channels, including short separator channels, reached acceptable or excellent levels for both quality indicators. Data were acquired at a 4.65 Hz sampling rate.

**Figure 1.**
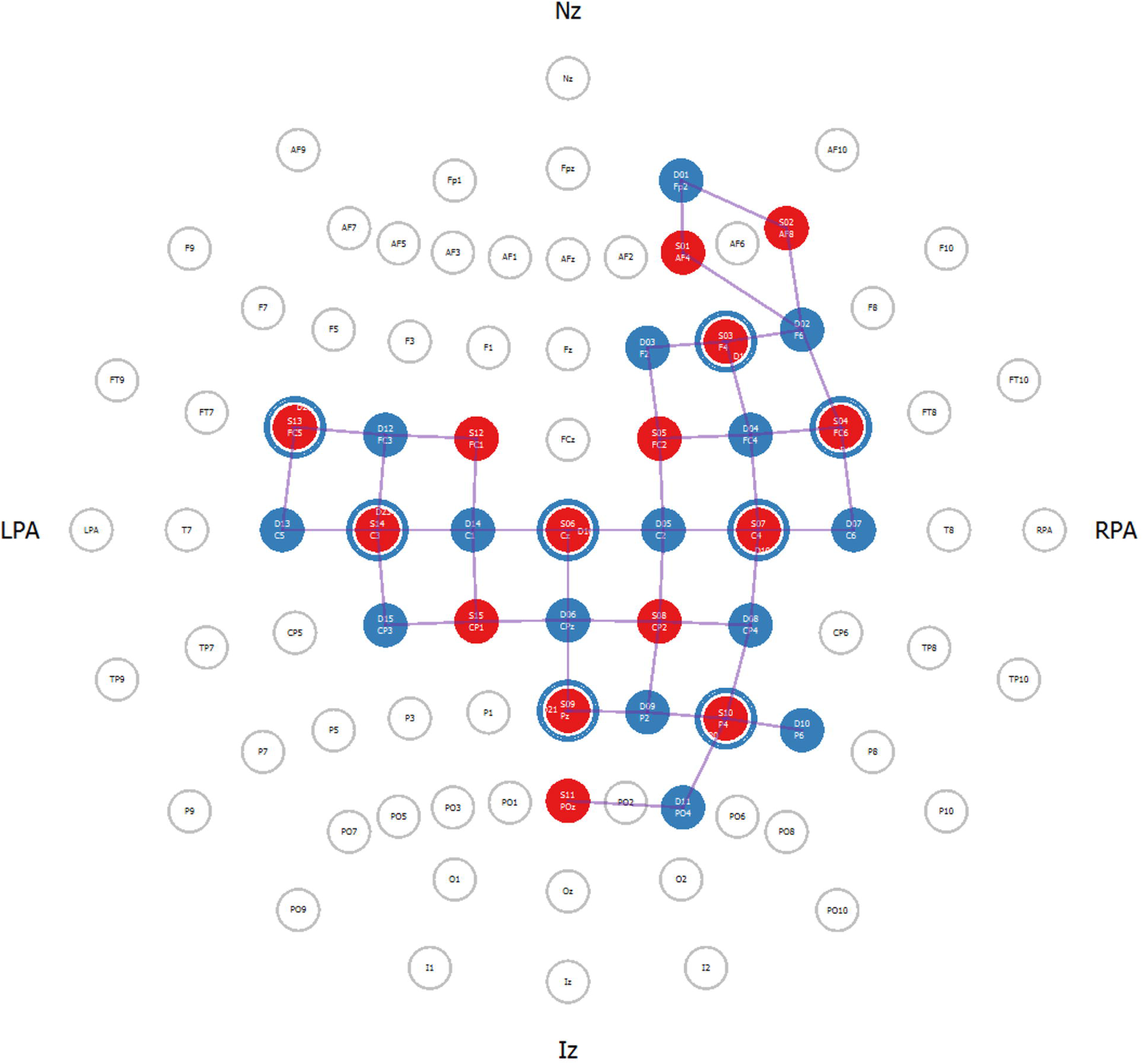
Functional near-infrared spectroscopy (fNIRS) head probe. *A priori* established regions of interest (ROI) for the fNIRS head probe. For this study, 15 LED sources and 15 detectors were used to create 50 channels covering the right lateralized nodes of the frontoparietal attention network and over bilateral primary motor and primary sensory cortices. Additionally, eight short-separator channels (illustrated as blue rings around red circles) were distributed throughout the head probe to measure scalp perfusion. Anatomical reference points are: nasion (Nz), right pre-auricular (RPA), inion (Iz) and left pre-auricular (LPA).

**Table 1.** FNIRS head probe – location of channels, sources, and detectors.

| Channel | Source | Detector | Channel coordinates (MNI) | AAL atlas location |
| --- | --- | --- | --- | --- |
| 1 | 1 | 1 | 25 58 10 | Frontal_Sup_R |
| 2 | 1 | 2 | 37 50 14 | Frontal_Mid_R |
| 3 | 2 | 1 | 34 63 0 | Frontal_Mid_R |
| 4 | 2 | 2 | 44 46 2 | Frontal_Inf_Tri_R |
| 5 | 3 | 2 | 42 34 19 | Frontal_Mid_R |
| 6 | 3 | 3 | 32 47 43 | Frontal_Mid_R |
| 7 | 3 | 4 | 41 20 30 | Frontal_Inf_Tri_R |
| 8 | 3 | 17 | 34 30 27 | Frontal_Mid_R |
| 9 | 4 | 2 | 46 24 19 | Frontal_Inf_Tri_R |
| 10 | 4 | 4 | 63 20 36 | Frontal_Inf_Oper_R |
| 11 | 4 | 7 | 65 6 23 | Precentral_R |
| 12 | 4 | 18 | 68 15 24 | Precentral_R |
| 13 | 5 | 3 | 18 24 45 | Frontal_Sup_R |
| 14 | 5 | 4 | 35 15 51 | Frontal_Mid_R |
| 15 | 5 | 5 | 22 -2 57 | Frontal_Sup_R |
| 16 | 6 | 5 | 15 -17 69 | Frontal_Sup_R |
| 17 | 6 | 6 | 1 -29 84 | Paracentral_Lobule_L |
| 18 | 6 | 14 | -13 -15 74 | Paracentral_Lobule_L |
| 19 | 6 | 16 | 1 -14 74 | Paracentral_Lobule_L |
| 20 | 7 | 4 | 53 0 49 | Frontal_Mid_R |
| 21 | 7 | 5 | 35 -16 52 | Precentral_R |
| 22 | 7 | 7 | 58 -11 43 | Postcentral_R |
| 23 | 7 | 8 | 36 -25 46 | Postcentral_R |
| 24 | 7 | 19 | 49 -18 48 | Postcentral_R |
| 25 | 8 | 5 | 32 -31 77 | Postcentral_R |
| 26 | 8 | 6 | 18 -47 78 | Parietal_Sup_R |
| 27 | 8 | 8 | 32 -42 51 | Parietal_Inf_R |
| 28 | 8 | 9 | 33 -56 72 | Parietal_Sup_R |
| 29 | 9 | 6 | 3 -43 58 | Precuneus_L |
| 30 | 9 | 9 | 15 -60 60 | Parietal_Sup_R |
| 31 | 9 | 21 | -2 -66 67 | Precuneus_L |
| 32 | 10 | 8 | 48 -53 50 | Angular_R |
| 33 | 10 | 9 | 36 -62 51 | Parietal_Sup_R |
| 34 | 10 | 10 | 41 -56 30 | Angular_R |
| 35 | 10 | 11 | 42 -79 42 | Occipital_Mid_R |
| 36 | 10 | 20 | 31 -59 43 | Angular_R |
| 37 | 11 | 11 | 21 -81 36 | Occipital_Sup_R |
| 38 | 12 | 12 | -29 12 54 | Frontal_Mid_L |
| 39 | 12 | 14 | -24 -1 64 | Frontal_Sup_L |
| 40 | 13 | 12 | -36 12 26 | Frontal_Inf_Oper_L |
| 41 | 13 | 13 | -38 6 23 | Frontal_Inf_Oper_L |
| 42 | 13 | 22 | -53 17 26 | Frontal_Inf_Tri_L |
| 43 | 14 | 12 | -32 -3 43 | Precentral_L |
| 44 | 14 | 13 | -48 -14 39 | Parietal_Inf_L |
| 45 | 14 | 14 | -26 -16 51 | Precentral_L |
| 46 | 14 | 15 | -42 -27 47 | Parietal_Inf_L |
| 47 | 14 | 23 | -43 -11 50 | Postcentral_L |
| 48 | 15 | 6 | -12 -43 78 | Postcentral_L |
| 49 | 15 | 14 | -17 -28 62 | Postcentral_L |
| 50 | 15 | 15 | -32 -42 58 | Parietal_Sup_L |

Channels, created by sources and detectors, were localized with Montreal Neurological Institute (MNI) coordinates and anatomical landmarks, as identified with AAL atlas. Short-separator channels are listed in blue font.

### FNIRS data pre-processing

FNIRS data files were processed using a proprietary software tool, Satori (v. 1.8) by Brain Innovation (NIRx Medical Technologies, nirx.net/satori). Satori includes a Graphical User Interface (GUI) where pre-processing steps can be selected; steps included motion artifact removal, physiological noise detection, and channel selection and removal. In Satori, preprocessing steps of conversion and spatial registration were performed automatically. Using the Modified Beer-Lambert Law,^5^ light intensity data were converted to optical density values and then to HbO, HbR and total hemoglobin (HbT) values. Additionally, data were spatially registered to our head probe and displayed for visual inspection and confirmation. Next, event markers files were manually created for each subject by renaming the event markers that are generated by PsychoPy. For the NC-DTS paradigm, PsychoPy sent numerical trial markers to Aurora via a lab streaming layer that corresponded with LE and UE subtask conditions. Next, the GUI was used to complete temporal pre-processing steps. Here, the Satori default parameters (10 interactions, 5s lag, 3.5 threshold, 0.5 influence) were used to complete motion artifact detection and correction with a spike removal procedure. When spikes were detected, a monotonic interpolation was applied, and then the Temporal Derivative Distribution Repair (TDDR)^29^ was applied to restore high frequency bands. Three steps were used to remove physiological noise. First, short separation regression (SSR) was completed through a generalized linear model (GLM), which uses the highest correlation method to select channels with artifacts automatically. Next, temporal filtering was completed to remove low-frequency drifts as well as portions of the non-hemodynamic related signal components (e.g. heart rate) using a Butterworth high-pass filter and then a Gaussian low-pass smoothing filter with cut-off frequencies of 0.01 Hz and 0.4 Hz, respectively. Finally, we applied a normalization step using the Z-Transform to make our data comparable across participants. We did not use the default setting in Satori that automatically rejects channels with a scalp coupling index (SCI) below 0.75^30^; this indicates that data originating from the light sources was poorly correlated. Instead, we processed all data twice – once with automated channel rejection and once without automated channel rejection. The files without automated channel rejection were used for group-level analysis, but the files that were processed with channel rejection were inspected to see which channels would have been rejected and their SCI values. This information was used after group-level analysis to check our data for potential outliers. After all these steps were completed, we used a multi-subject GLM approach in Satori to generate group-level data.

### FNIRS data analysis

Satori generated separate output files for the LE and UE subtasks. These files included averaged HbO, HbR, and HbT (not analyzed) beta weights for each participant, at each channel, and for each condition (i.e., dual, single motor, and single cognitive). The HbO beta weights were then organized into clusters of channels corresponding to the *a priori* functional ROIs. The channels associated with each cluster are as follows: for the right dorsolateral prefrontal cortex (PFC) cluster, the channels included were 1, 2, 3, 4, 5, 7, and 9; for the bilateral primary motor cortex (M1) cluster, the channels included were 16, 18, 20, 21, 43, and 45; for the bilateral primary somatosensory cortex (S1) cluster, the channels included were 22, 23, 26, 27, 44, 46, 48, and 50; and for the right posterior parietal cortex (PPC) cluster, the channels included were 28, 29, 30, 32, 33, and 34. An inherent limitation of traditional fNIRS montages is that each channel may cover multiple regions of interest due to the required space between source-detector pairs. Accordingly, the channels included in each cluster were selected using a combination of the AAL-2, Juelich, and Brodmann’s atlases in fOLD^25^ to maximize the likelihood of using channels that represent a specific functional ROI, while avoiding channels that overlap multiple functional ROIs.^31,32^

### Statistical analysis

Behavioral performance metrics and HbO beta weight clusters were then imported into RStudio (v. 2024.04.2, R version: 4.4.0) for statistical analysis. For both behavioral subtasks, a repeated measures analysis of covariance (RM ANCOVA) was computed, with group (repeat SRC or control), task condition, and sex as factors, and age and time since last SRC as covariates, to compare averaged dual task condition performance to averaged single task condition performance. Additionally, additional RM ANCOVA models were computed to test for differences in dual task effects (DTE)^33^ (see calculation below) between the control group and repeat SRC group, as this is a better performance metric because it accounts for within-subject variability on single task performance.

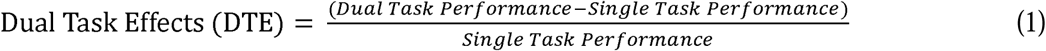

For the fNIRS functional ROI clusters, a repeated measures multivariate analysis of covariance (RM MANCOVA) was computed for the LE and UE subtask with group (repeat SRC or control), task condition, and sex as factors, and age and time since last SRC as covariates. Each of the four functional ROI clusters (PFC, M1, S1, and PPC) were included as separate variables. This was to assess if any of the factors or covariates had significant main effects or interactions at the whole-brain level. This is particularly relevant when the dependent variables (i.e., the functional ROI clusters) are likely to be correlated, as is often the case in neuroimaging data. Individual RM ANCOVA models were subsequently computed for each functional ROI cluster of each subtask with the same factors and covariates to assess the main effect and interaction for each cluster individually.

All RM ANCOVA and RM MANCOVA models used in this study met the necessary assumption requirements. Specifically, the assumption of independence of observations was upheld, with each subject’s data treated as independent from others. Normality was assessed by inspecting the distribution of residuals for each group across time points through examination of Q-Q plots. Homogeneity of variance was verified using Mauchly’s test of sphericity and Levene’s test. Linearity was confirmed through inspection of scatterplots, as recommended.^34^ Additionally, the assumption of homogeneity of regression slopes was confirmed by examining interactions between covariates and the categorical independent variable. For the MANCOVA models, multicollinearity among covariates was assessed by examining the correlation matrix to ensure no excessive correlation among independent variables. Post hoc pairwise comparisons were then calculated using paired t-tests for within-group comparisons and two sample homoscedastic t-tests for between-group comparisons. All t-tests were two-tailed with an alpha level threshold of *P* < 0.05 to determine significance. *P*-values were adjusted for multiple comparisons using false discovery rate (FDR), accounting for all combinations of behavioral comparisons and functional ROI comparisons. Additionally, Cohen’s *d* was used to calculate effect sizes for significant results.

### Data availability

The data that support the findings of this study are available from the corresponding author upon request. The code used for the analyses in this publication is available on GitHub at https://github.com/andyhagen11/Altered-neural-recruitment-despite-dual-task-performance-recovery-in-athletes-with-repeat-concussion

## Results

### Participants

A total of 37 participants were included in the analyses (20 with repeat SRC, 17 control). The mean age for the repeat SRC group was 20.9 years (SD: 1.49; Female: *n* = 11) and for control was 20.2 years (SD: 1.41; Female: *n* = 11). All participants were actively engaged in organized sports for at least four days per week. For the repeat SRC group, the median number of diagnosed SRCs was 3 (range: 2-5), and the mean time since the last diagnosed SRC was 1.75 years (SD: 1.90).

### Behavioral Data

#### Dual Task Performance

Behavioral data from the NC-DTS was analysed for both the raw performance of the single motor and dual tasks as well as the DTE for each participant. LE subtask: gait speed was slower for the dual vs. single task conditions (task main effect *F*(6.31), *P* = 0.014) but was not different between the repeat SRC and control groups (group main effect *F*(0.0024), *P* = 0.96). The task x group interaction was not significant (*F*(0.0021), *P* = 0.96). Additionally, there was no significant effect of age or sex on gait speed. UE subtask: number of catches was fewer in dual vs. single task conditions (*F*(13.27), *P* < 0.001) but was similar between groups (*F*(1.30), *P* = 0.26), and the task x group interaction was not significant (*F*(0.82), *P* = 0.37). Additionally, there was no effect of age, but a main effect of sex on task performance (*F*(5.78), *P* = 0.019), with males having better single motor task performance (*P* = 0.008, *d* = 0.90). Overall, our pairwise results indicate that for the pooled sample there was a difference between single and dual task for both gait (*P* < 0.001, *d* = 0.62) and catching performance (*P* < 0.001, *d* = 1.07) but the repeat SRC and control groups performed similarly (Figure 2).

**Figure 2.**
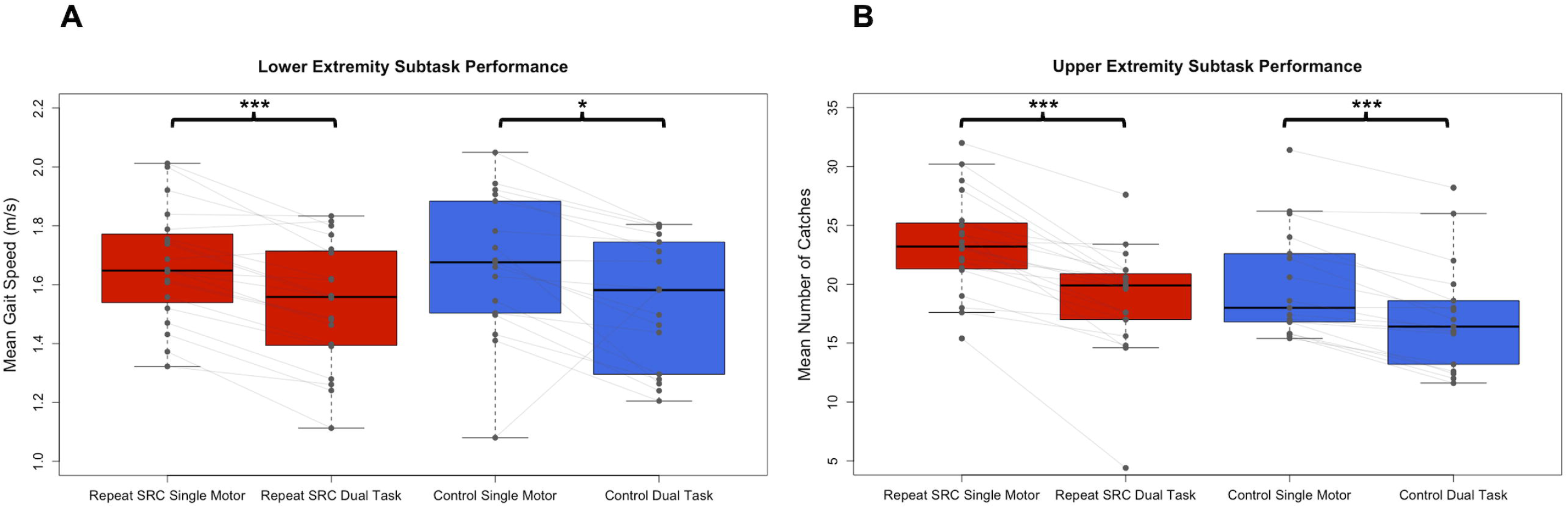
Dual task performance in the repeat SRC and control groups. (**A**) Mean gait speed (m/s) during the lower extremity (LE) single motor and dual tasks for the repeat SRC and control groups. (**B**) Mean number of catches during the upper extremity (UE) single motor and dual tasks for the repeat SRC and control groups. Light grey lines indicate paired single task and dual task performance for each group. All *P*s < 0.001 except for LE single motor to dual task for controls (*P* = 0.016).

#### Dual task effects (DTE)

DTE assesses the relative individual percent change from single to dual task performance, by quantifying both a motor DTE and a cognitive DTE.^33^ Consistent with the task performance results, there was a significant motor and cognitive DTE for both the LE subtask and UE subtask across the entire sample (LE motor DTE: *P* < 0.001, *d* = 0.68; LE cognitive DTE: *P* = 0.0027, *d* = 0.53; UE motor DTE: *P* < 0.001, *d* = 1.31; UE Cog DTE: *P* < 0.001, *d* = 0.61).

However, there was no main effect of group (*F*(1.04), *P* = 0.41) or pairwise differences in DTE between the repeat SRC and control groups (LE motor DTE: *P* = 0.70, *d* = 0.12; LE cognitive DTE: *P* = 0.055, *d* = 0.66; UE motor DTE: *P* = 0.17, *d* = 0.48; UE Cog DTE: *P* = 0.87, *d* = 0.05). Furthermore, neither age nor sex significantly affected any DTE, with the exception of a marginally significant effect of sex (*F*(4.24), *P* = 0.048) on the UE motor DTE, where females had less DTE (*P* = 0.035, *d* = 0.69). Figure 3 illustrates the motor DTE and cognitive DTE plotted against each other for both the LE and UE subtasks, which quantifies dual task interference.^35^ This data demonstrated no apparent prioritization of motor performance over cognitive performance or vice versa for the repeat SRC and control groups. Most participants experienced mutual inference of motor and cognitive performance (lower left quadrant), with a portion of participants experiencing a cognitive priority trade off (lower right quadrant).

**Figure 3.**
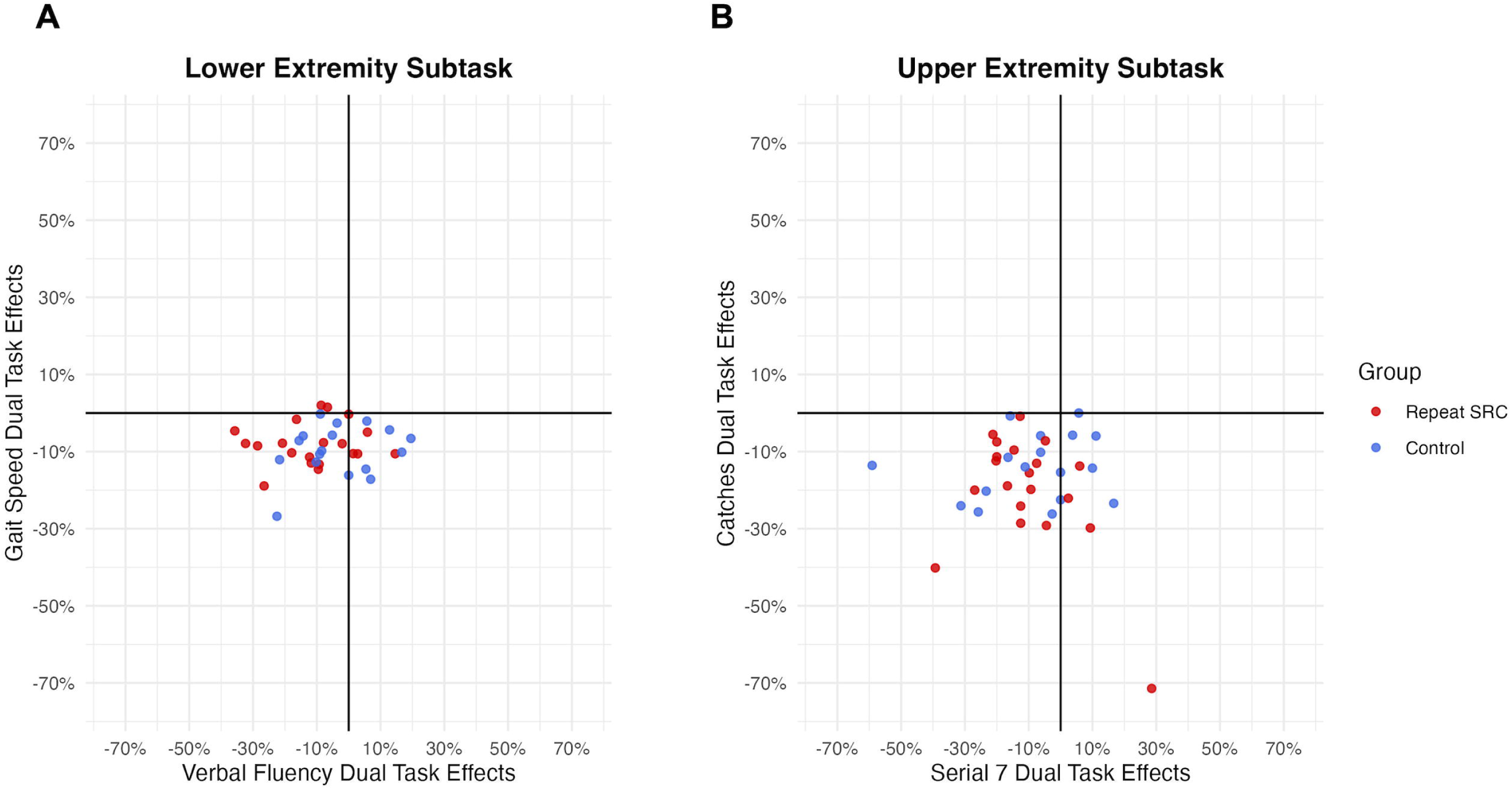
Dual task effects (DTE) and interference. **(A)** Individual motor (gait speed) DTE plotted against cognitive (verbal fluency) DTE for the lower extremity (LE) subtask. **(B)** Individual motor (catches) DTE plotted against cognitive (serial sevens) DTE for the upper extremity (UE) subtask. For both LE and UE, most participants experienced mutual inference of motor and cognitive performance (lower left quadrant), with a portion of participants experiencing a cognitive priority trade off (lower right quadrant).

**Figure 4.**
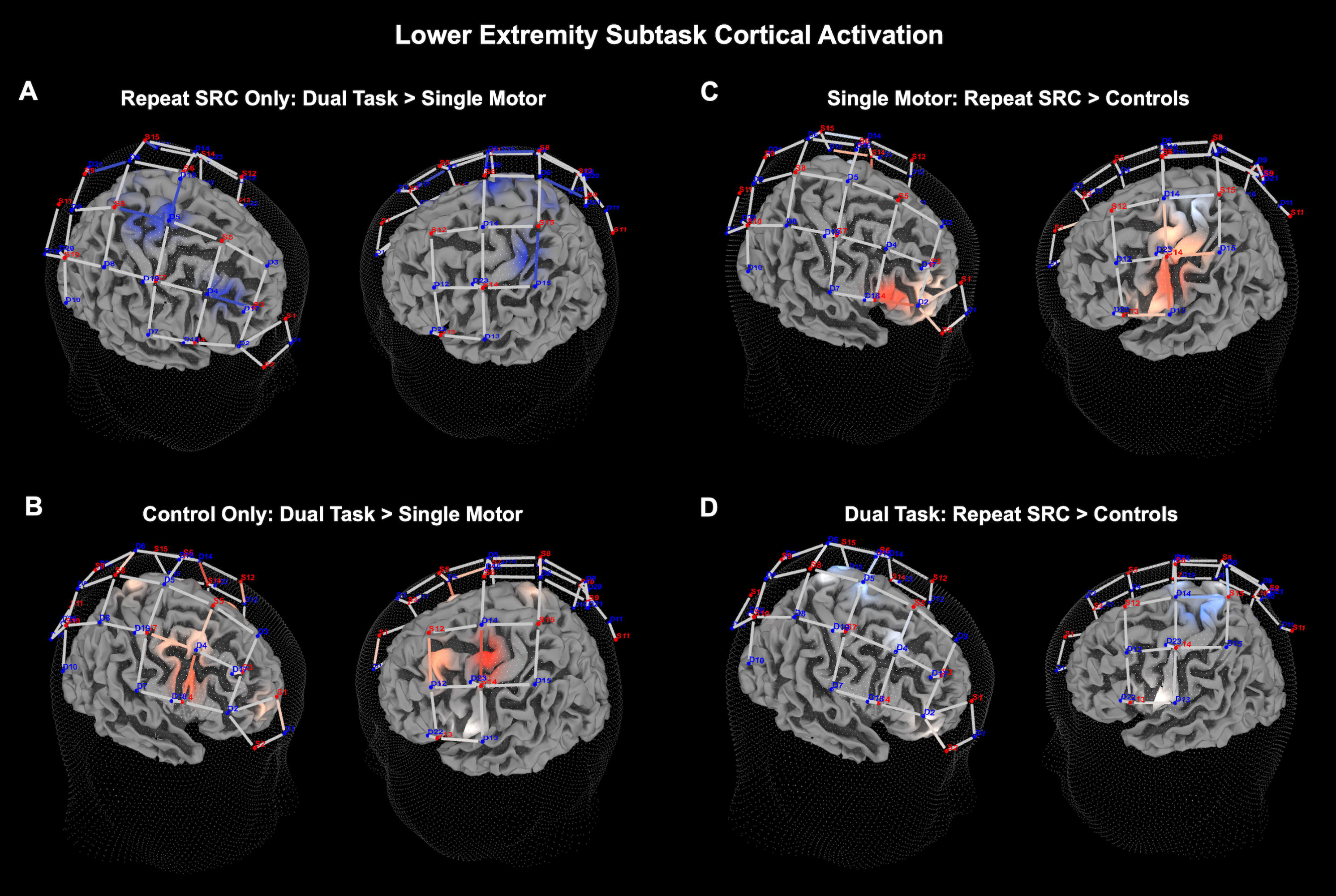
Lower extremity (LE) neural recruitment significance map. **(A)** Dual task activation contrasted against single motor task activation for the repeat SRC group only. **(B)** Dual task activation contrasted against single motor task activation for the control group only. **(C)** Activation in the repeat SRC group contrasted against activation of the control group for the single motor task condition. **(D)** Activation in the repeat SRC group contrasted against activation of the control group for the dual task condition. The colored channels illustrate the locations where significantly different activation was observed, and the light intensity indicates the magnitude of difference. Specifically, deeper shades of red indicate a greater magnitude increase in activation and deeper shades of blue indicate a greater magnitude decrease in activation.

**Figure 5.**
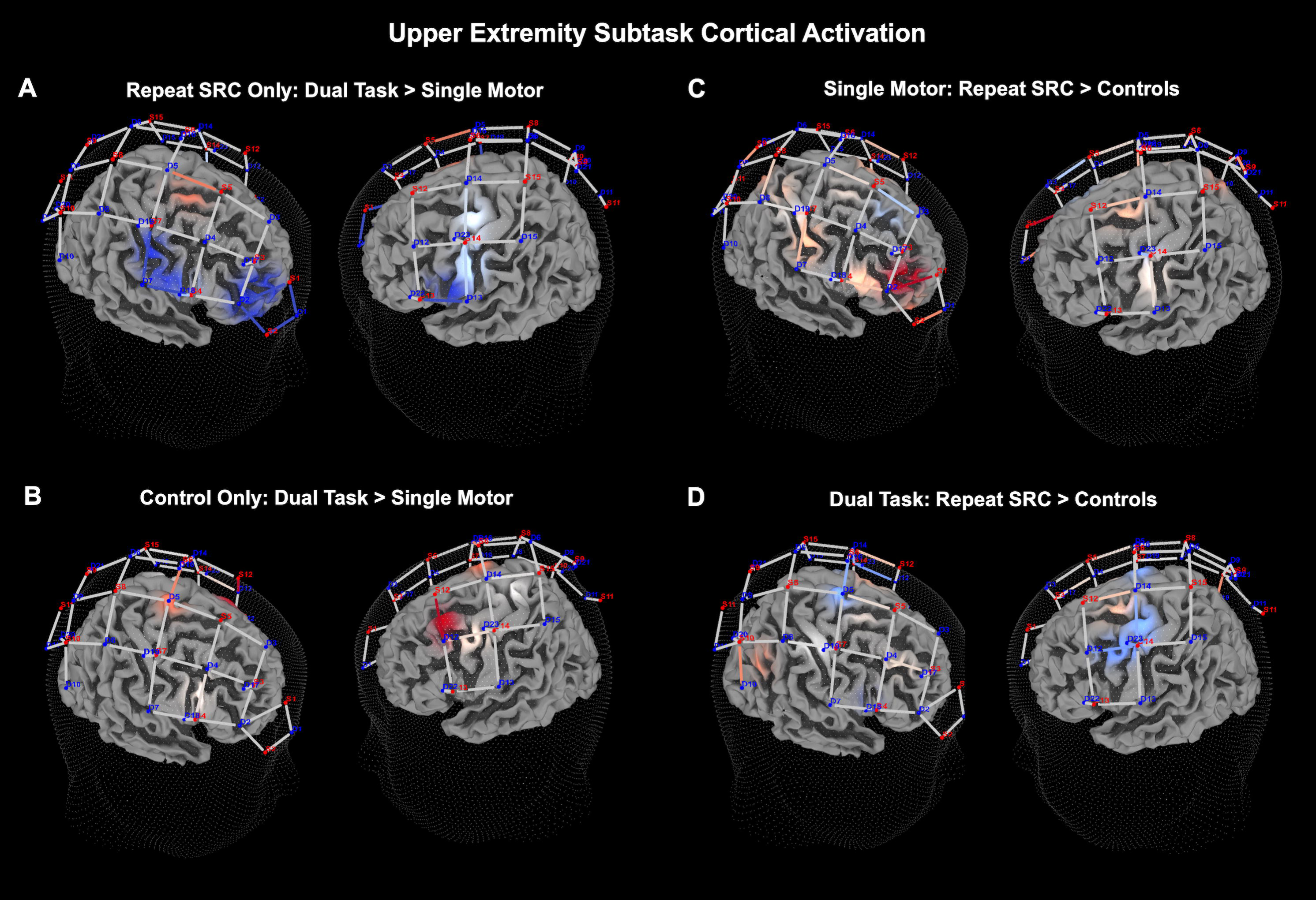
Upper extremity (LE) neural recruitment significance map. **(A)** Dual task activation contrasted against single motor task activation for the repeat SRC group only. **(B)** Dual task activation contrasted against single motor task activation for the control group only. **(C)** Activation in the repeat SRC group contrasted against activation of the control group for the single motor task condition. **(D)** Activation in the repeat SRC group contrasted against activation of the control group for the dual task condition. The colored channels illustrate the locations where significantly different activation was observed, and the light intensity indicates the magnitude of difference. Specifically, deeper shades of red indicate a greater magnitude increase in activation and deeper shades of blue indicate a greater magnitude decrease in activation.

### Task-Dependent Neural Recruitment

#### Lower extremity (LE) subtask fNIRS results

For the fNIRS outcomes, the channels were analysed as functional ROI clusters, including clusters for PFC, M1, S1, and PPC. Main effects of these analyses are described here, and pairwise comparisons are described in the following paragraph. The LE subtask RM MANCOVA revealed a significant main effect for task condition (*F*(3.32), *P* < 0.001), a significant main effect between the repeat SRC and control groups (*F*(3.01), *P* = 0.018), and a significant interaction between task condition and group (*F*(2.16), *P* < 0.028), indicating differences between task condition and group at the whole-brain level. Additionally, there were no significant effects of sex or age on the dependent variables. Examining individual RMANCOVA models for each functional ROI cluster, there were significant main effects of task condition for only the PFC cluster (*F*(14.2), *P* < 0.001), and significant main effects of group for only the PFC cluster (*F*(5.12), *P* = 0.024), and the PPC cluster (*F*(4.17), *P* = 0.042). For the interaction between task condition and group, there was significance in the PFC cluster (*F*(3.69), *P* = 0.025), the M1 cluster (*F*(4.35), *P* = 0.013), the S1 cluster (*F*(5.17), *P* = 0.0058), but not the PPC cluster. Furthermore, there was no effect of age or sex on any of the individual function ROI clusters.

Pairwise results demonstrated significant findings for the differences between task condition and group among the functional ROI clusters. Comparing dual task to single motor task activity, the repeat SRC group had slightly reduced activation in the M1 cluster (*P* = 0.038, *d* = 0.16), and the S1 cluster (*P* < 0.001, *d* = 0.23), with an absence of changes in the PFC or PPC clusters. Conversely, the control group had increased activation during the dual task in the PFC cluster (*P* = 0.0028, *d* = 0.19), the M1 cluster (*P* = 0.027, *d* = 0.15), and the S1 cluster (*P* = 0.025, *d* = 0.14) relative to single motor activation. Comparing the activation of the repeat SRC group to the control group during the single motor task, the repeat SRC group had increased activation in the PFC cluster (*P* = 0.0028, *d* = 0.40), with a lack of changes in the other functional ROI clusters. However, when comparing the repeat SRC group to the control group during the dual task, there were no significant changes in activation at the functional ROI cluster levels between groups. Comparing the activation of the repeat SRC group to the control group during the dual task, there were no significant changes in activation at the functional ROI cluster level between groups.

### Upper extremity (UE) subtask fNIRS results

For the UE subtask RM MANCOVA, with each individual ROI cluster as a dependent variable, there was a significant main effect between the repeat SRC and control groups (*F*(12.44), *P* < 0.001), but no significant main effect of task condition (*F*(1.07), *P* = 0.39), or an interaction effect (*F*(0.96), *P* = 0.46), likely attributable to neural recruitment across differing functional ROIs being correlated at the individual level. Additionally, there was no significant effect of sex or age on the dependent variables. Examining individual RMANCOVA models for each functional ROI cluster, there was a significant main effect of task condition for only the PFC cluster (*F*(3.85), *P* = 0.022), and significant main effects of group for the PFC cluster (*F*(13.49), *P* < 0.001), the M1 cluster (*F*(21.89), *P* < 0.001), the S1 cluster (*F*(4.37), *P* = 0.037), and the PPC cluster (*F*(10.55), *P* = 0.0012). For the interaction between task condition and group, there were no significant interactions for any functional ROI cluster. As with the RM MANCOVA results, these findings indicate significant changes in activation between the repeat SRC and control group, but a lack of effects of task condition or an interaction effect. Furthermore, there was no effect of age or sex on any of the individual function ROI clusters.

Considering these findings, there were significant pairwise differences between groups among the functional ROI clusters. Comparing dual task to single motor task activity, the repeat SRC group had a significantly reduced activation in the PFC cluster (*P* < 0.001, *d* = 0.29), with an absence of changes in the M1, S1, and PPC clusters. Conversely, the control group had increased activation during the dual task condition in only the M1 cluster (*P* = 0.0052, *d* = 0.20). Comparing the activation of the repeat SRC group to the control group during the single motor task, the repeat SRC group had substantially increased activation in the PFC cluster (*P* < 0.001, *d* = 0.47), with a lack of changes in the other functional ROI clusters, which is congruent with the LE subtask findings. Finally, comparing the activation of the repeat SRC group to the control group during the dual task, there was a significant decrease in activation for the repeat SRC group in the M1 cluster (*P* = 0.0052, *d* = 0.42) and no other significant differences at the cluster-level.

## Discussion

This study aimed to investigate the impact of repeat SRCs on dual task performance and its associated neural recruitment in college-aged athletes. Using functional near-infrared spectroscopy (fNIRS), we measured neural recruitment in the frontoparietal attention network and bilateral primary motor and sensory cortices while athletes with repeat SRCs and control athletes performed the NC-DTS.^21^ The results suggest that athletes with repeat SRCs have comparable dual task performance relative to controls, yet they exhibit chronically altered neural recruitment strategies to support this behavioral performance.

### Athletes with repeat concussion have typical dual task performance

The results of this study strongly suggest that athletes with a history of repeat SRCs have dual task performance that is comparable to control athletes who have never experienced an SRC. Although we didn’t monitor these athletes longitudinally, their typical performance probably reflects that, if they had dual task deficits, those have recovered. While this finding may appear contrary to previous well-documented dual task impairments associated with SRC,^36^ our unique sample has a substantially longer time since their last SRC, with an average of 1.75 years since their last diagnosed SRC. While many studies have demonstrated dual task impairment following an SRC, even following symptom resolution and being cleared for return-to-play, most studies employed a sample who experienced an SRC much more recently and a sample that lacks repeat SRCs.^36^ The novelty of these results is that they are from an evaluation of longer-term effects of repeat SRCs rather than previously described acute effects. Other data has suggested that dual task performance often recovers within six months for most collegiate athletes, and more rapidly for females.^3,37^ This is congruent with our findings that most of the repeat SRC group had similar dual task performance and DTE to healthy controls. Interestingly, we also identified a slight effect of sex, with females having reduced UE motor DTE than males, which further strengthens the notion of sex differences in dual task recovery following SRC.^3,37^

### Increased PFC recruitment during single motor tasks in athletes with repeat concussion

The study revealed significantly elevated PFC activity in athletes with repeat SRCs during single motor task conditions compared to controls. This heighted neural recruitment in athletes with repeat SRCs was evident in both the LE single motor task and the UE single motor task, further demonstrating a consistent effect across multiple types of motor tasks. In agreement, other studies have shown an increase in PFC activation in a variety of different tasks^38,39^ and a greater recruitment of functional neural networks to support behavioral performance in athletes with SRC.^40^ This observed hyperactivation during both the LE and UE single motor tasks may reflect compensatory neural recruitment necessary to maintain behavioral performance levels in tasks that would typically require less cognitive input. Heightened PFC activation has also been reported during typical gait for those with persistent post-concussive symptoms and has suggested to be compensatory for a reduction in motor automaticity.^41^ This compensatory PFC recruitment in our sample may serve as a neural marker for the lingering effects of SRC. This suggests that even simple motor tasks like walking have reduced automaticity and demand greater cognitive control in athletes with repeat SRCs, and this extends beyond symptom resolution and medical clearance for return-to-play. It is not entirely clear if these changes signify brain recovery or ongoing impairment,^7^ as the neural recruitment patterns were not associated with our measured behavioral differences. However, these findings could be considered in the context of the compensation-related neural circuit hypothesis (CRUNCH), which describes neural activation patterns observed in aging adults who have reduced neural efficiency and structural brain changes.^42,43^ Specifically, increased PFC activity during single motor tasks – as seen here in young adults with repeat SRC – may reflect neural inefficiency, which could be a sign that these specific neurophysiological differences are reflective of some degree of ongoing impairment.

### Reduced PFC and M1 recruitment during dual tasks in athletes with repeat concussion

While there were few differences in activation between athletes with repeat SRCs and control athletes during dual task conditions, a closer examination of the intra-athlete changes from single motor to dual task conditions provides deeper insight into changing recruitment strategies. During dual task conditions, athletes with repeat SRCs displayed considerably reduced activation in PFC for the UE subtask and in M1 for the LE subtask, compared to single motor conditions. This response contrasts with the typical and well-documented dual task neural response characterized by increased activation in PFC and M1.^17,44^ The difference between the dual task neural recruitment changes for the UE and LE subtasks likely could be attributable to the novelty of the wall-toss task compared to the automaticity involved in gait. Nevertheless, both task paradigms generate significant dual task effects and are effective for assessing dual task performance, being sensitive in both upper and lower extremity motor control. Further, this finding is incongruent with the increased PFC activation during single motor tasks, suggesting a potential disengagement or overload of these regions during dual task processing. While DTE were similar between groups, this reduced activation may indicate a reduced capacity to engage the necessary cognitive and motor neural substrates simultaneously. This may result in poorer integration of dual task demands that impacts performance and injury risk at higher levels of task difficulty, such as during sports.

Previous research, using different imaging modalities, has also suggested that individuals who have a history of concussions and are asymptomatic have altered allocation of attentional resources. For instance, a study by Tapper et al.^45^ using EEG demonstrated that athletes with a history of SRC have deficits in sensory gating and cognitive processing, represented by reduced amplitudes of N1 and P3 event-related potentials during dual task performance. Further, other studies have shown that reduced amplitude of N2 and P3b potentials during a novelty oddball task can be detected long-term following a concussion, despite normative cognitive performance.^46^ Collectively, these findings, along with ours, suggest that there are ongoing changes in neurophysiology in individuals with repeat SRC. However, as stated above these changes may or may not reflect ongoing impairment,^8^ but this could be better delineated in future work by adding behavioral tasks with greater difficulty and sensitivity to subtle deficits.

Indeed, most lab-based dual task assessments likely do not emulate the same level of difficultly as dual task demands during sport performance, especially in the motor domain. While athletes with repeat SRCs can perform similarly to control athletes in lab-based assessments like ours, they may have increased dual task impairments during more challenging task conditions than presented in this study. Considering our findings that suggest neural compensation and disengagement in athletes with repeat SRCs, it is possible that this behavioral performance would not be sustained under more demanding task conditions. An alternative theory is that the atypical brain activity observed provides evidence of not just impairment or vulnerability, but also a form of recovery. This could signify that neural recruitment is adapting in a way that allows them to perform well in controlled settings, but it is unknown if this adaptation will suffice in high-demand, complex environments like sports. Considering the robust evidence linking increased rates of musculoskeletal injury to a previous SRC, often attributable to dual task impairments,^36^ our findings of chronic neural compensation and disengagement may play a significant role in these observations.

### Limitations

There were several limitations in this study that warrant consideration. The repeat SRC group sample had a large standard deviation in time since the athletes’ last SRC (1.90 years). While there was no relationship between time since last SRC and neural recruitment or behavioral performance in our sample, there are likely other neural changes that emerge over time that we did not measure. Further, the cross-sectional design of the study precludes causal inferences and limits the examination of long-term changes in neural recruitment and dual task performance. Future research should consider longitudinal studies with larger cohorts to confirm these findings and explore the trajectory of behavioral recovery and its relationship with neural recruitment over time, since it remains unclear whether observed changes in neural recruitment signify brain recovery or indicate persistent impairment. Additionally, while fNIRS provides valuable information about neural activation, specifically during real-world tasks such as walking, it has limitations in spatial resolution and depth of measurement compared with other functional neuroimaging techniques such as fMRI. The fNIRS head probe is only able to measure activation at the cortical surface and cannot assess subcortical and cerebellar contributions to performance on the NC-DTS.

### Conclusion

These findings highlight the relationship between behavioral recovery and neural compensation in athletes with a history of repeat SRCs. While these athletes may exhibit typical single task and dual task performance, this is accompanied by distinct neural recruitment alterations, including hyperactivation of the PFC during single motor tasks and reduced PFC and M1 activation during dual task conditions. The observed patterns of neural compensation and disengagement suggest altered utilization of neural resources that may prove unsustainable under more demanding conditions, potentially heightening the risk of future injury. These findings underscore the importance of considering both behavioral and neural outcomes when assessing recovery from SRCs. Greater understanding of compensatory neural recruitment strategies that support behavioral performance following repeat SRCs can better inform return-to-play decisions, future musculoskeletal injury risk, and the long-term impact of SRCs on neurocognitive function.

## Funding

This research was funded by the National Institutes of Health, grant numbers 5K01HD096047-02 and K12HD055931 issued to author J.S.

## Competing interests

The authors report no competing interests.

## Notes

### Competing Interest Statement

The authors have declared no competing interest.

### Author Declarations

The biomedical institutional review board of Colorado State University gave ethical approval for this work.

